# Risk Factors for 30-day Postoperative Surgical Site Hematoma Requiring Evacuation After Resection of Brain Metastases

**DOI:** 10.1101/2022.11.22.22282636

**Authors:** Yilong Zheng, Kejia Teo, Vincent Diong Weng Nga, Tseng Tsai Yeo, Mervyn Jun Rui Lim

## Abstract

**Objective:** To identify the risk factors for a 30-day postoperative surgical site hematoma requiring evacuation (POH) after surgical resection of brain metastases.

**Methods:** Patients who underwent surgical resection of brain metastases between 2011 and 2019 at our institution were included. Risk factors for a 30-day POH were identified using a multivariate logistic regression model.

**Results:** A total of 158 patients were included in the analysis. The mean (SD) age of the study population was 59.3 (12.0) years, and 82 (53.2%) patients were female. The incidence of a 30-day POH was 8.2% (13 patients). There was no statistically significant association between the occurrence of a 30-day POH and overall mortality (p=0.100). On multivariate analysis, there was a statistically significant association between a 30-day POH and younger age (OR=0.91; 95% CI=0.83, 0.99; p=0.035), higher BMI (OR=1.61; 95% CI=1.16, 2.46; p=0.010), and blood type AB (OR=21.7; 95% CI=1.66, 522; p=0.031). On receiver operating characteristic analysis, a threshold BMI of 25.1 kg/m^2^ and threshold age of 57 gave the optimum balance of sensitivity and specificity in predicting the occurrence of a 30-day POH.

**Conclusions:** Patients below 57 years old, who have a BMI of above 25, and/or have blood type AB were at higher risk of developing a 30-day POH after surgical resection of brain metastases. Additional care in intraoperative hemostasis and postoperative monitoring may be indicated among patients who have these risk factors.

## Introduction

Brain metastases are the commonest malignancies of the brain.^1,2^ Surgical resection is important in the management of brain metastases because it confers survival benefit,^3-6^ allows for symptomatic relief,^7^ and allows for a definitive diagnosis to be made.^7^ However, there are significant risks associated with surgical resection, such as a postoperative surgical site hematoma requiring evacuation (POH).

The incidence of POH specifically among patients with brain metastases is unclear. However, the incidence of POH after craniotomy for intracranial tumors in general has been reported to range from 1.8% to 2.1%,^8-10^ and risk factors include older age,^9,10^ higher blood international normalized ratio (INR),^9^ larger tumor size,^9^ and infratentorial location.^8^

Prior studies on the risk factors for POH included patients who underwent craniotomy for any type of intracranial tumor.^8-10^ While this constitutes important information, conclusions from these studies may not be generalizable to patients with brain metastases. Therefore, we aimed to identify the risk factors for a 30-day postoperative surgical site hematoma requiring evacuation after resection of brain metastases.

## Methods

### Study design

A retrospective study of patients who underwent surgical resection of brain metastases at our institution between March 2011 and December 2019 was conducted. Institutional ethics approval was obtained from the institutional review board prior to study initiation (National Healthcare Group Domain Specific Review Board; Reference Number 2020/00358). A waiver of informed consent was granted as this study posed no more than minimal risk to participants.

### Cohort selection

The operating theatre records database was accessed to retrieve the National Registration Identity Card numbers (NRICs, which act as the national identification number) of patients who underwent surgical resection of a brain tumor. The electronic medical records of the patients were then accessed and screened for inclusion in the study. Patients who (1) had a histologically verified metastasis to the brain, and (2) were 18 years old or above on the day of surgical resection were included.

### Data collection

Clinical data were collected using a standardized data collection form. Variables collected include (1) demographics, including age, sex, ethnicity, preoperative height, weight, body mass index (BMI), smoking history, (2) medical history, including hypertension, hyperlipidemia, diabetes mellitus, history of stroke or transient ischemic attack, (3) details on the primary cancer, including presence of extracranial metastases preoperatively and site of the primary tumor (defined as the 5 commonest sites and others), (4) details on the brain metastasis, including presence of preoperative intratumoral hemorrhage (as reported by the radiologist in the radiology report), volume of the largest resected tumor on preoperative MRI (defined as AP x ML x CC/2, where AP, ML, and CC denote the greatest anteroposterior, medial-lateral, and craniocaudal diameters), presence of preoperative hydrocephalus, extent of midline shift, extent of resection (gross total or subtotal resection), presence of residual tumor on the first postoperative MRI as reported by the radiologist, (5) results of the preoperative full-blood count closest to the surgical resection, including white blood cell count, red blood cell count, hemoglobin level, mean corpuscular volume, hematocrit, platelet count, neutrophil count, lymphocyte count, monocyte count, eosinophil count, basophil count, (6) results of the preoperative clotting studies closest to the surgical resection, including the prothrombin time (PT), activated partial thromboplastin time (aPTT), INR, and (7) the ABO blood type of the patient.

### Statistical analysis

The baseline characteristics of the patients were reported using mean and standard deviation for continuous variables and count numbers and percentages for categorical variables. For hypothesis testing against the occurrence of a 30-day POH, the Pearson’s χ2 test was used for categorical variables, and the student’s t-test was used for continuous variables. The Fisher’s exact test was used for variables with a sample size of lesser than 5 under any category. Statistical significance was defined as a p-value of 0.050 or lower.

Variables with a p-value of lesser than 0.200 on univariate analysis against the occurrence of a 30-day POH were included as independent variables in a multivariate logistic regression model, with the occurrence of a 30-day POH as the dependent variable. The independent variables that had a p-value of 0.050 or lower on multivariate logistic regression analysis were considered as risk factors for a 30-day POH. For the risk factors that were continuous variables, the threshold value that optimally discriminated a 30-day POH occurrence from non-occurrence was derived using receiver operating characteristic analysis.

To determine whether a 30-day POH was associated with overall mortality, univariate time-to-event analysis using the Kaplan-Meier method was performed. Hypothesis testing was performed using the log-rank test, and a p-value of 0.050 or lower was taken to be statistically significant. For patients who died during the follow-up period, time-to-mortality was defined as the duration between the first surgical resection of the brain metastases and the date of death. For patients who did not die during the follow-up period, time-to-mortality was defined as the duration between the first surgical resection of the brain metastases and the date of the latest follow-up. All data analyses were conducted using R Studio Version 1.2.5042.

## Results

### Baseline characteristics of the study population

The baseline characteristics of the study population were reported in Table 1. A total of 158 patients were included in the analysis. The mean (SD) age of the study population was 59.3 (12.0) years, and 82 (53.2%) patients were female. The incidence of a 30-day POH was 8.2% (13 patients). The median overall survival of patients with and without a 30-day POH were 11.3 and 12.4 months respectively. There was no statistically significant association between the occurrence of a 30-day POH and overall mortality (p=0.100) (Figure 1).

**Figure 1:** Kaplan-Meier curve evaluating the association between occurrence of a 30-day postoperative surgical site hematoma requiring evacuation and overall mortality Abbreviation: POH, postoperative surgical site hematoma requiring evacuation.

**Table 1:** Baseline characteristics of patients with and without a 30-day postoperative surgical site hematoma requiring evacuation

| Variables | No 30-day POH<br>(n=145, 91.8%) | 30-day POH<br>(n=13, 8.2%) | Total (n=158) | p-value |
| --- | --- | --- | --- | --- |
| Age; mean (SD) | 59.8 (12.0) | 54.1 (10.4) | 59.3 (12.0) | 0.099 |
| Ethnicity; n (%) |  |  |  | 0.207 |
| Chinese | 100 (69.0) | 7 (53.8) | 107 (67.7) |  |
| Malay | 22 (15.2) | 2 (15.4) | 24 (15.2) |  |
| Indian | 7 (4.8) | 0 (0.0) | 7 (4.4) |  |
| Others | 16 (11.0) | 4 (30.8) | 20 (12.7) |  |
| Female; n (%) | 76 (52.4) | 8 (61.5) | 84 (53.2) | 0.575 |
| Height; mean (SD) | 160.9 (9.8) | 158.3 (9.3) | 160.8 (9.8) | 0.410 |
| Weight; mean (SD) | 58.2 (13.2) | 65.3 (17.0) | 58.8 (13.6) | 0.072 |
| Body Mass Index (kg/m<sup>2</sup>); mean (SD) | 22.4 (4.2) | 27.2 (6.8) | 22.7 (4.5) | **0.001** |
| Smoking history; n (%) |  |  |  | 0.789 |
| Never smoker | 96 (67.6) | 9 (81.8) | 105 (68.6) |  |
| Ex-smoker | 30 (21.1) | 1 (9.1) | 31 (20.3) |  |
| Current smoker | 16 (11.3) | 1 (9.1) | 17 (11.1) |  |
| On antiplatelets preoperatively; n (%) | 14 (9.7) | 1 (7.7) | 15 (9.5) | 1.000 |
| On anticoagulants preoperatively; n (%) | 0 (0.0) | 1 (7.7) | 1 (0.6) | 0.082 |
| Hypertension; n (%) | 48 (33.1) | 5 (38.5) | 53 (33.5) | 0.762 |
| Hyperlipidemia; n (%) | 36 (24.8) | 1 (7.7) | 37 (23.4) | 0.303 |
| Diabetes mellitus; n (%) | 24 (16.6) | 4 (30.8) | 28 (17.7) | 0.249 |
| History of stroke or TIA; n (%) | 3 (2.1) | 0 (0.0) | 3 (1.9) | 1.000 |
| Presence of extracranial metastasis preoperatively; n (%) | 95 (65.5) | 5 (38.5) | 100 (63.3) | 0.071 |
| Site of primary tumor; n (%) |  |  |  | 0.664 |
| Lung | 57 (39.3) | 6 (46.2) | 63 (39.9) |  |
| Breast | 36 (24.8) | 4 (30.8) | 40 (25.3) |  |
| Colorectum | 18 (12.4) | 0 (0.0) | 18 (11.4) |  |
| Skin | 7 (4.8) | 0 (0.0) | 7 (4.4) |  |
| Kidney | 5 (3.4) | 1 (7.7) | 6 (3.8) |  |
| Others | 22 (15.2) | 2 (15.4) | 24 (15.2) |  |
| Preoperative intratumoral hemorrhage; n (%) | 33 (22.8) | 7 (53.8) | 40 (25.3) | **0.021** |
| Volume of the largest resected tumor; mean (SD) | 22.2 (35.6) | 16 (18.5) | 21.7 (34.5) | 0.534 |
| Preoperative hydrocephalus; n (%) | 31 (21.4) | 2 (15.4) | 33 (20.9) | 1.000 |
| Preoperative midline shift (mm); mean (SD) | 3 (4.5) | 1.2 (2.4) | 2.9 (4.4) | 0.166 |
| Gross total resection; n (%) | 113 (77.9) | 9 (69.2) | 122 (77.2) | 0.495 |
| Residual tumor postoperatively; n (%) | 36 (24.8) | 5 (38.5) | 41 (25.9) | 0.324 |
| Preoperative full blood count; mean (SD) |  |  |  |  |
| White blood cell count | 10.1 (3.7) | 8.9 (3.3) | 10 (3.7) | 0.257 |
| Red blood cell count | 4.5 (0.7) | 4.3 (0.7) | 4.4 (0.7) | 0.438 |
| Hemoglobin | 12.8 (1.7) | 12.5 (2.4) | 12.8 (1.8) | 0.623 |
| Mean corpuscular volume | 87.1 (7.5) | 87 (6.2) | 87.1 (7.4) | 0.950 |
| Hematocrit | 38.4 (4.8) | 36.7 (7.2) | 38.3 (5.1) | 0.244 |
| Platelet count | 257.2 (83.3) | 225.4 (83.0) | 254.6 (83.5) | 0.186 |
| Neutrophil count | 8.8 (6.0) | 7.1 (3.6) | 8.7 (5.8) | 0.313 |
| Lymphocyte count | 1.5 (1.6) | 1.2 (0.7) | 1.5 (1.6) | 0.493 |
| Monocyte count | 0.6 (0.6) | 0.5 (0.2) | 0.6 (0.5) | 0.335 |
| Eosinophil count | 0.1 (0.2) | 0.1 (0.2) | 0.1 (0.2) | 0.666 |
| Basophil count | 0.0 (0.0) | 0.0 (0.0) | 0.0 (0.0) | 0.930 |
| Preoperative clotting studies; mean (SD) |  |  |  |  |
| PT | 12.1 (1.4) | 12 (1.7) | 12.1 (1.4) | 0.717 |
| aPTT | 26.6 (3.2) | 26.4 (2.8) | 26.6 (3.2) | 0.875 |
| INR | 1.2 (1.9) | 1.0 (0.1) | 1.2 (1.8) | 0.783 |
| Blood type |  |  |  | **0.010** |
| AB | 9 (6.2) | 5 (38.5) | 14 (8.9) |  |
| O | 52 (35.9) | 3 (23.1) | 55 (34.8) |  |
| B | 41 (28.3) | 2 (15.4) | 43 (27.2) |  |
| A | 43 (29.7) | 3 (23.1) | 46 (29.1) |  |

Abbreviations: POH, postoperative surgical site hematoma requiring evacuation; SD, standard deviation; BMI, body mass index; TIA, transient ischemic attack; PT, prothrombin time; activated partial thromboplastin time; INR, international nationalized ratio.

### Risk factors for a 30-day POH

On multivariate analysis, younger age (OR=0.91; 95% CI=0.83, 0.99; p=0.035), higher BMI (OR=1.61; 95% CI=1.16, 2.46; p=0.010), and blood type AB (OR=21.7; 95% CI=1.66, 552; p=0.031) were independently associated with a higher risk of a 30-day POH (Table 2).

**Table 2:** Multiple logistic regression model for the identification of risk factors for a 30-day postoperative surgical site hematoma requiring evacuation

| Variables | OR | 95% CI | p-value |
| --- | --- | --- | --- |
| Age | 0.91 | 0.83, 0.99 | **0.035** |
| Weight | 0.90 | 0.78, 1.01 | 0.116 |
| Body Mass Index | 1.61 | 1.16, 2.46 | **0.010** |
| Presence of extracranial metastasis preoperatively | 0.33 | 0.06, 1.71 | 0.188 |
| Intratumoral hemorrhage on preoperative MRI |  |  |  |
| No | — | — |  |
| Yes | 1.37 | 0.14, 11.0 | 0.774 |
| Preoperative platelet count | 0.99 | 0.97, 1.00 | 0.123 |
| Blood type |  |  |  |
| A | — | — |  |
| AB | 21.7 | 1.66, 552 | **0.031** |
| B | 1.49 | 0.10, 26.3 | 0.770 |
| O | 1.30 | 0.11, 16.8 | 0.828 |

Abbreviations: OR, odds ratio; CI, confidence interval.

On receiver operating characteristic analysis, a threshold BMI of 25.1 kg/m^2^ gave the optimum balance of sensitivity (80.0%) and specificity (76.6%) in predicting the occurrence of a 30-day POH (AUC=0.74; 95% CI=0.56, 0.92). On the other hand, a threshold age of 57 gave the optimum balance of sensitivity (61.5%) and specificity (66.2%) in predicting the occurrence of a POH (AUC=0.66; 95% CI=0.52, 0.79).

## Discussion

In our cohort of patients who underwent surgical resection of brain metastases, younger age, higher BMI, and blood type AB were found to be risk factors for a 30-day POH. Specifically, a threshold age of 57 and threshold BMI of 25.1 provided the optimum balance of sensitivity and specificity in predicting the occurrence of a 30-day POH.

Prior studies reported that older age was associated with a higher risk of a POH.^9,10^ However, in our cohort, younger age was associated with a higher risk of a POH. The higher risk of POH among older patients has been attributed to greater tissue fragility among older patients.^10,11^ The association between younger age and higher risk of POH in our cohort may be due to the fact that we generally aim for a greater extent of resection for younger patients, so as to reduce the likelihood of recurrence within their lifetime. However, a greater extent of resection also carries with itself a theoretically higher risk of damage to the surrounding vasculature, thereby increasing the risk of a POH.

In our cohort, a higher BMI was associated with a higher risk of POH as well. However, prior studies for other types of brain tumors found no statistically significant association.^12,13^ The association between higher BMI and POH may be attributed to the fact that patients with higher BMI are also more likely to have comorbidities that predispose them to POH, such as hypertension. However, our supplementary analyses revealed no statistically significant association between BMI and a history of hypertension (p=0.556). Further studies evaluating the association between BMI and POH among patients with brain metastases are needed.

Our analyses also revealed blood type AB to be an independent risk factor for a 30-day POH. Existing studies on the association between blood type and bleeding diathesis reported that patients with blood type O had the highest risk of bleeding, and this association may or may not be related to lower levels of von Willebrand factor and factor VIII among patients with blood type O.^14^ On the basis of these findings, it follows logically that in our cohort, patients with blood type O should have a higher risk of a 30-day POH. However, blood type AB, not O, was associated with a higher risk of a 30-day POH in our cohort. The association between blood type and POH in our cohort may be spurious, especially considering the relatively small sample size (only 14 patients had blood type AB) and the relatively wide 95% confidence interval for the association between blood type AB and the occurrence of a 30-day POH (95% CI=1.66, 552). However, the possibility of ABO blood type having an actual effect on the risk of a 30-day POH is not ruled out. Further studies clarifying the association between blood type and POH among patients with brain metastases are needed.

Our study has several limitations. First, as this was a single-center study, our conclusions may not be generalizable to other institutions. Second, the relatively small sample size of our cohort limited the statistical power of our analysis. Future studies of a similar nature should aim to include the data of multiple centers to ensure the generalizability of conclusions and overcome the issues associated with a small sample size.

## Conclusions

Patients below 57 years old, who have a BMI of above 25, and/or have blood type AB were at higher risk of developing a 30-day POH after surgical resection of brain metastases. Additional care in intraoperative hemostasis and postoperative monitoring may be indicated among patients who have these risk factors.

## Data Availability

All data produced in the present study are available upon reasonable request to the authors

## Acknowledgements

None.

